# TASTE identifies shared proteomic effects on multiple related cancers

**DOI:** 10.64898/2025.12.19.25342717

**Authors:** Debepsita Mukherjee, Kevin Wang, Andrew J. Song, Stella Koutros, Ludmila Prokunina-Olsson, Mark Purdue, Stephen Chanock, Mitchell Machiela, Diptavo Dutta, Indranil Sahoo

## Abstract

**Introduction:** Genome-wide association studies (GWAS) have identified hundreds of variants linked to cancers, but their downstream regulatory consequences remain poorly understood. Increasing evidence suggests that related cancers share alterations of common regulatory programs. Trans-associations of cancer risk variants mediated via molecular phenotypes, such as gene expression and protein levels, can help uncover these downstream mechanisms. Further investigation of such convergence can reveal shared etiological processes and therapeutic targets across cancer types.

**Method:** We introduce TASTE (Trans Association using Shared factorization and TEsting), a summary statistic-based framework to identify protein sets that are trans-regulated by genetic variants associated with sets of biologically related cancers. TASTE consists of three steps: (1) **TASTE-D**, a low-rank matrix factorization to estimate shared and group-specific trans-association patterns across cancers; (2) **TASTE-S**, a sparse singular value decomposition to identify proteins driving shared effects; (3) **TASTE-T**, a competitive testing strategy for evaluating significance of trans-associations captured by the identified protein-set.

**Results:** Simulation studies show that TASTE can identify protein-set with shared effects across groups of related cancers with high accuracy. Applying TASTE to UK Biobank plasma proteomic data and GWAS of genitourinary cancers (renal cell carcinoma [RCC] and bladder cancer [BLCA]), we identified 88 proteins with shared *trans*-associations. Notably, 43% of these proteins lacked significant trans-pQTL but were discovered through aggregation of weak signals only. This includes **KDR** (p=1.4×10^-7^), a VEGF receptor and drug target for RCC and BLCA, nominally associated with 10 RCC and 8 BLCA loci. Additional *in silico* analyses identified enrichment in cancer-relevant pathways such as PI3K/AKT, MAPK, VEGF, and Ras signaling, as well as significant protein-protein interaction and differential expression in tumors. Identified proteins were overrepresented in known cancer drivers and genes essential to cell proliferation in RCC and BLCA cell lines.

**Conclusion:** TASTE is a robust and scalable approach to uncover convergent trans-regulatory mechanisms across related cancers or diseases enabling discovery of downstream molecular targets and highlighting candidates for mechanistic follow-up and therapeutic development.

## Introduction

Genome-wide association studies (GWAS) have identified tens of thousands of common variants associated with complex traits and diseases^1^, including about 30 cancers and sub-types. A majority of these identified trait-related variants map to the non-coding region of the genome^2^. To understand their broader biological functions, studies have subsequently integrated data on intermediate molecular phenotypes, such as gene expressions, protein levels, and metabolites^3–6^. This has been facilitated by the development of high-throughput technologies to profile such molecular omics and emergence of several large-scale omics studies^3,6–10^. In particular, the studies of the plasma proteome have increasingly garnered attention due to the potential to investigate a wide range of biological processes, some showing potential for drug targeting^6,11,12^. Through integrative analysis of proteogenetic data, several studies have demonstrated a noticeable overlap of the disease-related variants with protein quantitative trait loci (pQTL) i.e. variants associated with the protein levels and have identified target proteins potentially associated with diseases, especially cancers ^9,10,13,14^.

The majority of investigation of GWAS loci till date have focused on identifying the effects of genetic variants on the regulation of physically proximal proteins (cis-associations/cis-effects)^6,11^. However, recent studies show that a higher proportion of disease heritability is mediated via the association of disease-related variants with distal molecular targets (trans-associations/trans-effects)^15^. The proposed “omnigenic” model suggests that the effects of the genetic variants associated with complex diseases, including cancers, propagates through intricate regulatory networks and converge on a smaller set of disease-relevant ‘core’ molecular targets or proteins^16^. If such an “omnigenic” characterization is reflective of the genetic architecture of diseases, then the trans-pQTL associations of disease-related variants could identify such “core” proteins, to which effects of multiple genetic variants (usually single nucleotide polymorphisms; SNPs) potentially converge and thus highlight key regulatory mechanisms underlying disease etiology^17^. However, detecting trans-pQTL effects remains challenging due to confounding effects^18^, weaker effects of SNPs on distal proteins compared to cis-regions, resulting in reduced statistical power, and an increased multiple testing burden^19^. This has led to development of innovative statistical methods to identify trans-associations, especially for disease-related variants, converging on a smaller group of molecular targets^20–22^.

So far there has been a notable overlap between molecular targets (genes, proteins and others) and common biological processes associated across related diseases, for example related cancers. For example, PI3K/AKT/mTOR pathway and vascular endothelial growth factor pathways are commonly dysregulated in urogenital cancers (renal, bladder and prostate cancers)^23–27^. This has led to the development and consideration of mTOR inhibitors and anti-angiogenic therapies for these malignancies^28–30^. Additional notable examples include altered expressions of BCL2 and MYC in several hematological malignancies^31–33^ and regulation of Wnt/β-Catenin pathway in digestive system cancers^34,35^. Thus, such observed overlap in underlying mechanisms can be leveraged to identify shared proteomic effects. This could lead to insights into shared biological mechanisms and inform future therapeutic strategies^36,37^.

Here we present a summary statistic-based method Trans Association using Shared factorization and <u>TE</u>sting (TASTE) to identify proteins that have shared trans-associations with genetic variants associated to related diseases. We focus on identifying shared proteomic effects across related cancers, and genetic variants associated to a particular cancer is considered a ‘group’. Using z-statistic for association between proteins and multiple groups of genetic variants corresponding to related cancers, TASTE performs three broad steps (**Figure 1**): first TASTE-D, we perform a low-rank decomposition of the association matrix across multiple groups, to estimate the shared effect sizes and disease-specific effect sizes between variants and proteins. Then for TASTE-E, we perform a sparse singular value decomposition to identify sets of proteins which exhibit strong contributions in the shared effects. Finally, in TASTE-T we perform a competitive testing^38^ to evaluate whether the selected sets of proteins explain an enriched proportion of variant-protein associations, against a broader background of polygenic characteristics. Through simulations, we show that TASTE consistently identifies the shared effects with high accuracy under a number of settings. We apply TASTE to analyze two categories of related cancers: (1) blood/hematological cancers including chronic lymphocytic leukemia, multiple myeloma, Hodgkin’s lymphoma and (2) genitourinary cancers: kidney and bladder, using data from the current large-scale cancer GWAS, and the UK Biobank pharma proteomics project (UKB-PPP)^10^ for 2,940 plasma proteins. We identified sets of 161 and 88 proteins with significant shared trans­associations with variants related to cancers in the two groups respectively. In several downstream in silico analyses, we show that the identified proteins have evidence to be relevant for multiple cancers.

**Figure 1:**
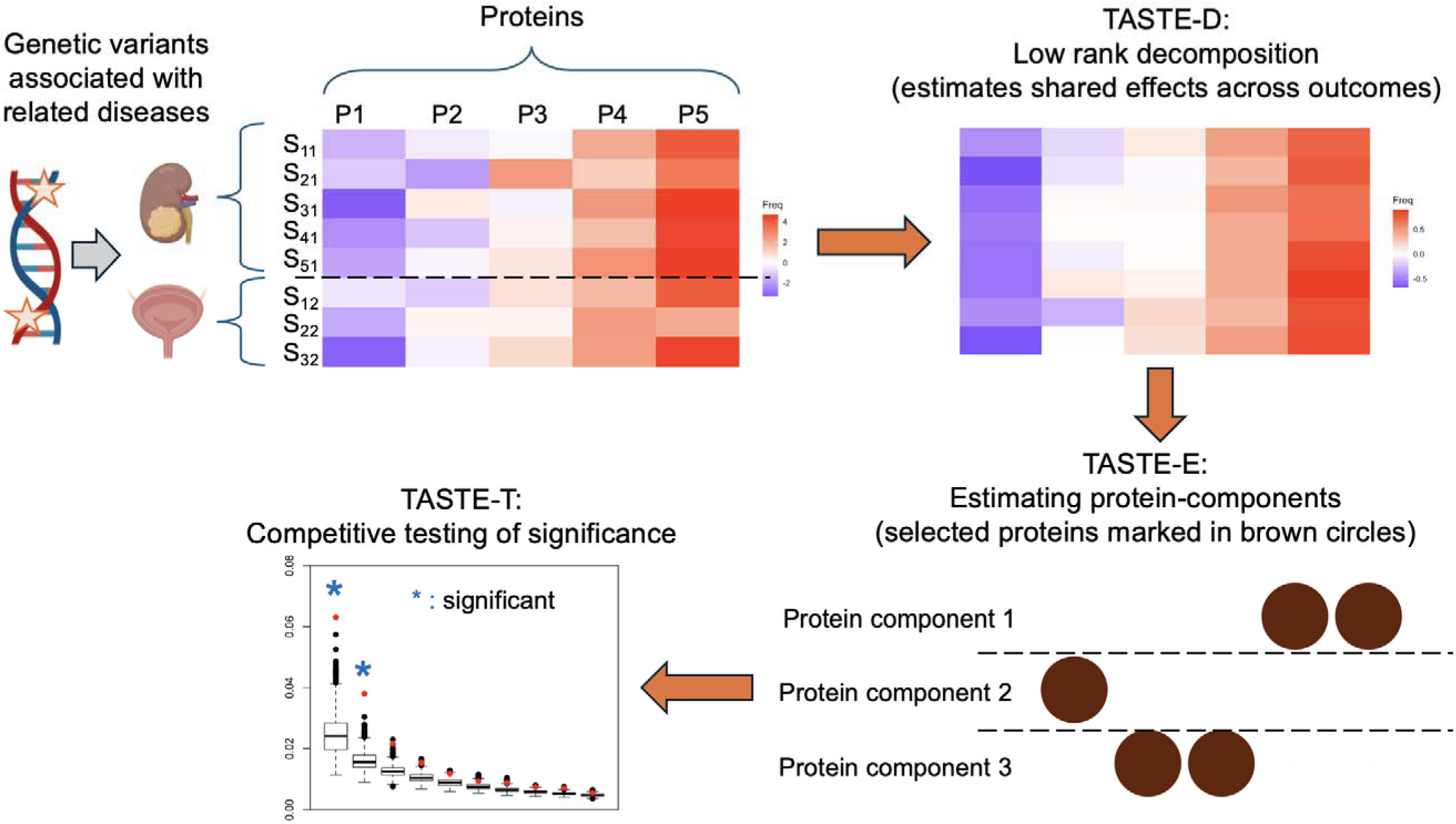
Overview of TASTE. A toy example to illustrate TASTE. Using z-statistic for association between proteins and multiple groups of genetic variants corresponding to related cancers TASTE: (1) TASTE-D a low-rank decomposition to estimate the shared effect sizes (2) TASTE-E, a sparse singular value decomposition to identify sets of proteins (protein components) which exhibit strong contributions in the shared effects (selected proteins are marked in brown circles) and (3) TASTE-T: a competitive testing to evaluate the significance of the identified protein-sets.

## Materials and Methods

We consider *c* groups of genetic variants, which in our context correspond to *c* related cancers. For each cancer type, a set of independent significant SNPs can be identified through existing large-scale GWAS or from public resources like PGS-catalog^39^. For the *k*^th^ cancer, we assume that summary statistics data (*Z* values and *p* values) from standard trans-pQTL analysis are available for *g_k_* GWAS significant SNPs across *p* (>> *g_k_*) proteins that are distal to the SNPs. Using this information, we construct a data matrix *Z_k_ (g_k_ × p)* of trans-pQTL summary statistics for each group (cancer), where *Z_k;i,j_* represents the *Z*-value of association between the í^th^ SNP and the /^h^ protein for *k* = 1,…,c. Starting with the summary statistics matrices for each cancer (or group), TASTE has three broad steps.

### Step 1. TASTE-D: Low rank Decomposition

We augment the summary statistics matrices *Z_k_* to form a single data matrix *Z* as:

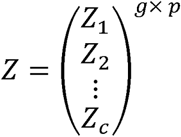

where 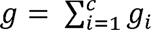. The primary aim of this step is to decompose the augmented data matrix across *c* cancers, into a low-rank matrix of joint or shared effects *(**R**)* and a matrix of cancer-specific effects (***S***). There are several available methods for such decomposition such as Multivariate Analysis of Variance (MANOVA), Joint and Individual Variation Explained (JIVE)^40^, Multi-Omics Factor Analysis (MOFA)^41^, Multi-set Correlation and Factor Analysis (MCFA)^42^ among others, all of which can estimate *R.* In this article, we use the JIVE algorithm which additively decomposes *Z* as:

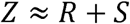

where *R,* is the low rank matrix of rank *r* for the estimated shared effects given by

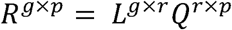

where *Q^r^*^×^*^p^* is a matrix to map *p* proteins on a latent space of rank *r* while *L^g×r^* is the contribution of genetic variants to that space, and

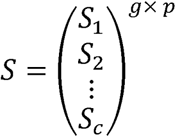

where *S_i_* is the matrix of cancer specific effects. Although the above algorithms/methods can broadly be used for matrix decomposition, these have typically been developed for exploratory analyses of individual-level data. Here, we tailor them to use summary statistics from omics studies, specifically pQTLs.

### Step 2. TASTE-E: Identifying protein-components using sparse singular value decomposition

In this step, we identify protein sets which have the largest contributions in the shared effect matrix ***R***, through sparse singular value decomposition. We estimate a sparse linear vector ***v*** such that:

*(v) = argmax **v**^T^**R**^T^**Rv***

with 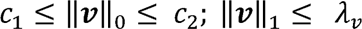 and ||***v***||_2_ = 1. Here ***v*** denotes a sparse loading vector, corresponding to columns of ***R***, i.e. the proteins (**v** is termed ‘protein component’ subsequently). Each non-zero element of the protein-component (**v**) indicates that the respective protein is selected. Intuitively, the selected proteins represent sets that have trans-associations with the selected SNPs, serving as potential mediators of the SNPs’ effects (See **Supplementary Methods** for a detailed algorithm). We define a corresponding s-value for protein component (**v**) as the square of the largest eigen value of ***R****^T^**R***. It can be interpreted as the strength of the contribution of the selected proteins in the shared effect matrix.

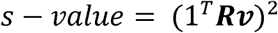

This metric is a measure of the cumulative association captured by the proteins through aggregating multiple (possibly weaker) associations. Here, 1 denotes a vector of length *p*, containing 1’s.

We can estimate *r* such protein sets where the rank of ***R*** is *r*, using matrix deflation, analogous to singular vectors of a matrix. Each such subsequently estimated protein component can be interpreted as distinct biological mechanisms, due to the orthogonality between the components. We devised an estimation algorithm for protein components such that there is no overlap in terms of the proteins selected in these components, imposing strict orthogonality. See **Supplementary Methods** for a detailed algorithm.

### Step 3. TASTE-T: Testing of significance of protein components in a competitive framework

To assess which protein-components significantly capture the group-specific *trans­*association patterns, we evaluate the results from the original analysis against a *competitive* null hypothesis. Since GWAS significant variants are expected to be enriched for *trans-*pQTLs in general, we test whether the s-values obtained in the original analysis are higher than those obtained using the *trans-*pQTL summary statistics between a randomly selected set of GWAS-identified variants and proteins of similar size, that, by design, do not exhibit any group-specific patterns. For this, we first construct a *null matrix* by randomly sampling *g* variants from the pool of all independent GWAS significant variants and extracting the corresponding *trans-*summary statistics for another set of randomly chosen *p* genes. This matrix of *trans-*associations, by design, should not reflect group-specific patterns.

We then apply TASTE-D and TASTE-E to extract the protein components and calculate corresponding s-values. We repeat this step multiple *(M)* times to generate a competitive null distribution of s-values. To assess statistical significance, we compare the observed s-values from the original analysis against their respective null distributions to compute empirical p-values. Specifically, the p-value for the k^th^ protein component is given by

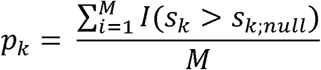

where *s_k_* is the observed s-value for the *k*^th^ protein component and *s_k_*_;*null*_ is a percentile of the corresponding null distribution.

To summarize, TASTE identifies proteins which have potentially shared effects across the *c* groups, by first identifying the shared effects of all proteins across groups (TASTE­D), then selecting the protein-sets (via protein components) having the highest contribution to the shared effects (TASTE-E) and finally, testing whether these protein­sets exhibit any group-specific patterns of association or whether these could have been identified using random group assignments (TASTE-T).

### Simulations

We carried out simulations to evaluate the performance of TASTE in identifying shared effects and nominating proteins through protein-components across several scenarios. We particularly focused on the accuracy of TASTE in estimating the rank of the shared effects matrix (TASTE-D) and accuracy (sensitivity and specificity) in identifying the proteins which have the highest contributions (TASTE-E).

We first generate a matrix 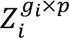 of z-values for the *í*^th^ group with *g_i_* SNPs and *p* proteins as follows:

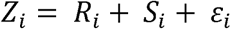

where *Ri* = *UiV*, *S_i_ = L_i_Q_i_* and *ε_i_* denotes an error matrix of independent gaussian observations. *U_i_* is a matrix of rank *r* and dimensions *g_i_ × r*, constructed by *g_i_* independent observations from a multivariate normal distribution, *MVN(0, I_r_). V* is a matrix of dimensions *r × p*, constructed by concatenating two matrices as: *V = [βI_r_*, 0*_r;p-r_*], where 0*_r;p-r_* is a matrix of 0 with *r* rows and *(p - r)* columns. Here, *β* denotes the effect for each of the first *r* proteins which are assumed to have a common effect across multiple groups. The matrix of specific effects *S_i_* of rank *r_i_* is constructed using two components. The first component *L_i_* is a *g_i_ × r_i_* matrix of rank *r_i_*, formed using *g_i_* independent observations from *MVN(0, I_ri_)*. The second component *Q_i_* is a *r_i_xp* matrix formed by setting randomly chosen *r_i_* columns to be *δI_r_*, while all remaining entries in the matrix are set to zero. We set *δ* = *β*/2, which denotes the effect for each of *r_i_* proteins which have group specific effects. In constructing *Q_i_*, we choose the *r_i_* columns such that they do not overlap with the proteins chosen to have shared effects across groups and do not overlap with the proteins chosen to have group specific effects for other groups. A detailed algorithm outlining the data generation process is described in **Supplementary Methods**.

Thus, the setting produces a matrix 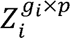 of z-values which has *r* proteins that have shared effects across multiple groups, each having effect *β,* and *r_i_* proteins that are group specific each having effect *δ*, while the remaining proteins are not associated. In our simulations, we evaluated the performance of TASTE using two groups (*i* = 2) under a total of 6 distinct scenarios with varying values of *g_i_ r* and *r_i_*.

## Results

### Simulation results

The results from the simulations are displayed in **Figure 2**. For both lower *(g*_1_ = 50 and *g*_2_ = 40) and higher *(g*_1_ = 150 and *g*_2_ = 100) number of genetic variants in each group, TASTE-D can reliably estimate the rank of the shared effects matrix, with the variation in estimates decreasing with joint rank *(r)* and common effect of shared proteins *(β),* as expected (**Figure 2A-B**). This indicates that our algorithm can identify the common patterns of variation across the two groups with the accuracy increasing with higher overlap between the groups and higher number of genetic variants in each group. Additionally, **Figures 2C-D** show that TASTE-E can identify the shared proteins accurately, with high sensitivity and specificity. Across all the settings, TASTE-E can identify the sets of proteins which have a shared effect across the groups while having a low number of “null” proteins, which are the proteins that do not have a shared effect. This is manifested in both the sensitivity and specificity of TASTE-E under multiple settings. As above, both the metrics increase with joint rank *(r)* and common effect of shared proteins (*β*) highlighting the improvement in performance of TASTE-E with higher overlap between the groups. However, though our simulation results are promising, they cannot possibly capture the complexity of genetic regulation of molecular phenotypes and only serve as broad performance metric in controlled scenarios. To our knowledge, no current method systematically addresses such joint estimation of proteomic (or molecular) effects across multiple groups and hence we lack a comprehensive benchmarking procedure for TASTE.

**Figure 2:**
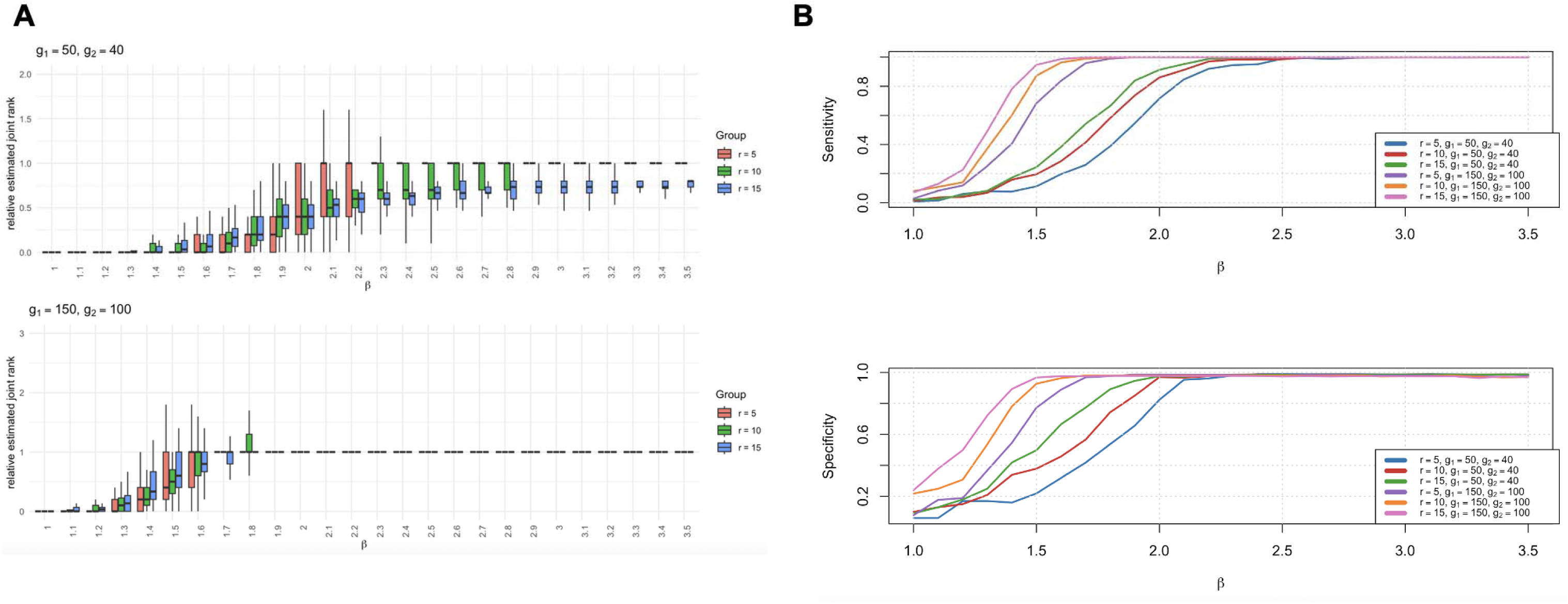
Simulation performance of TASTE. **(A)** Estimation of rank of shared matrix (joint rank) by TASTE-D across (upper panel) lower and (lower panel) higher number of genetic variants in each group. Vertical axis represents the ratio of the estimated rank and the true rank (*r*), colored by 3 different values of *r*. Horizontal axis represents signal strength () relative to noise for proteins “shared” across groups. **(B)** Sensitivity (upper panel) and Specificity (lower panel) of TASTE-E in identifying the proteins shared across the groups of va**ria**nts in the category

### Analysis of UK Biobank proteomics: Groups of related cancers

Next, we focus on results for two categories of related cancers and identify the corresponding selected target proteins using TASTE. For SNPs associated to each cancer, we constructed a z-value matrix *Z_i_* by extracting the Z-values for all the proteins whose transcription start site lies outside a +/- 1Mb window around the SNP. For each category, the set of proteins analyzed was obtained by taking the proteins common across all the cancers belonging to that category. For each category, we then applied TASTE to obtain the protein components reflecting the trans-association patterns for the corresponding category. Through several downstream analyses, we provide evidence that the identified proteins in the protein component have relevance to the overall genetic architecture of the respective cancers and thus merit further investigation as possible downstream targets underlying shared processes.

### Hematologic cancers

Hematologic cancers, encompassing leukemias, lymphomas, and multiple myeloma, collectively constitute approximately 9% of all new cancer diagnoses and 9.1% of all cancer-related deaths in the USA^43^. We focus on three hematological cancers, namely, chronic lymphocytic leukemia (CLL), multiple myeloma (MM), and Hodgkin’s lymphoma (HL), which have 41, 21, and 20 associated SNPs, respectively^44^. In total, our analysis considered 82 SNPs, and 2,711 proteins located distally (> 1Mb) from all of these SNPs. Using TASTE, we identify two significant components comprising a total of 161 proteins (91 in component 1, 70 in component 2) which potentially represent shared biological processes underlying these cancers (**Figure 3A**; **Supplementary Table 1)**.

**Figure 3:**
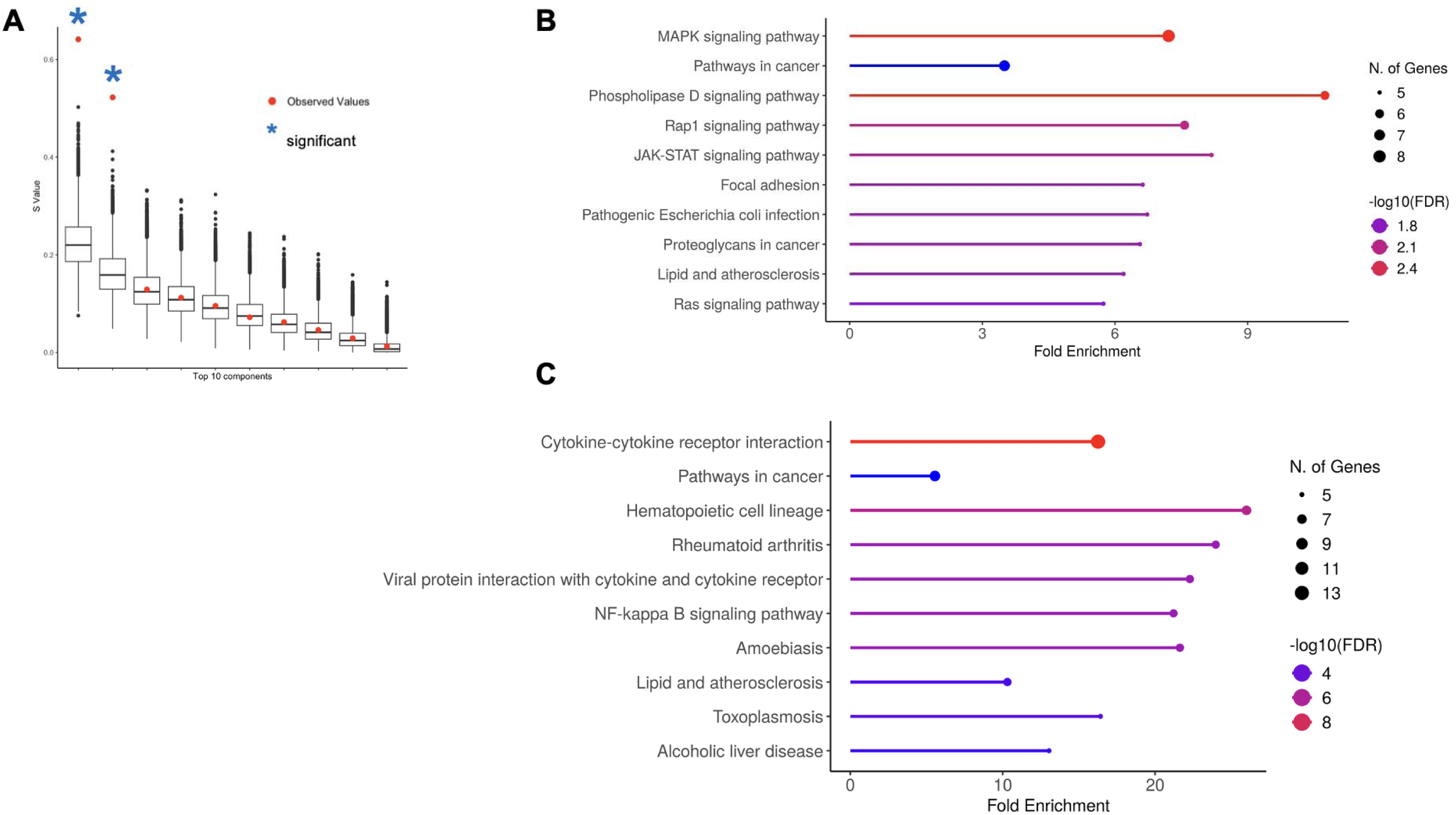
Results for blood/hematological cancers. **(A)** Observed s-values (a measure of overall trans-associations captured by the protein components identified by TASTE; See **Methods**) in red dots against the expected distribution of s-values using a competitive hypothesis for the top 10 protein components. The significant associations are marked in blue asterisk. Pathway analysis for **(B)** protein component 1 and **(C)** protein component 2 showing the fold enrichment, number of overlapping proteins, and -log_10_(FDR) for significance.

Among the 161 proteins identified, 74 did not have a strong trans-pQTL (p-value < 2.2×10^-07^: Bonferroni threshold for all pairwise comparisons with 82 SNPs and 2,711 proteins) association with any one of the 82 SNPs. Furthermore, 28 proteins had strong trans-pQTL associations in multiple cancers within the group of which TASTE identified 23, including known proteins like BCL2L1, indicating that the rest were identified by aggregating weaker trans-associations. Notably, this included proteins like AKT serine/threonine kinase 2 (AKT2; minimum p-value = 7.7×10^-04^) and diazepam binding inhibitor (DBI; minimum p-value = 2.1×10^-03^) which have been implicated in multiple hematological and lymphoid cancers^45^.

Further in silico analysis of TASTE components revealed that the 91 proteins selected in protein component 1 had significant physical protein-protein interactions (p-value < 1×10^-16^). This indicates that the proteins form important biological complexes potentially consequential to downstream disease manifestation. The selected proteins are also enriched in several known cancer processes like MAPK signaling pathway (FDR = 2.7×10^-03^), Rap1 signaling pathway (FDR = 9.1×10^-03^), and Ras signaling pathway (FDR = 0.02) among others (**Figure 3B**; **Supplementary Table 2**). Similarly, 70 proteins identified in protein component 2 are also enriched in protein-protein interactions (p-value < 1×10^-16^) and in known processes like Cytokine-cytokine receptor interaction (FDR = 0.01) and NF-kappa B signaling pathway (FDR = 0.02) among others (**Figure 3C**; **Supplementary Table 3**), which have been linked to immunoregulation. Overall, among the 161 proteins identified, 3 proteins are reported to be cancer drivers in COSMIC^46^ while 19 are candidate cancer drivers^47^ (**Supplementary Table 5**). These highlight the potential relevance of the identified proteins for hematological cancers as a group.

### Genitourinary cancers

Approximately one in four of all cancer cases and one in ten of all cancer deaths in USA are projected to be attributable to genitourinary cancers and are expected to increase owing to the aging population^43^. Here we consider two common genitourinary cancers: bladder cancer (BLCA) and renal cell carcinoma (RCC) which have 62 and 105 SNPs associated respectively^48,49^ reported in UKB-PPP data. Removing overlapped SNPs and proteins within +/- 1Mb of them, our analysis focused on a set of 167 SNPs and 2709 distal (trans-) proteins. TASTE identified one protein component to be significant comprising of 88 proteins in total broadly representing shared proteomic effects across these malignancies (**Figure 4A**; **Supplementary Table 1**).

**Figure 4:**
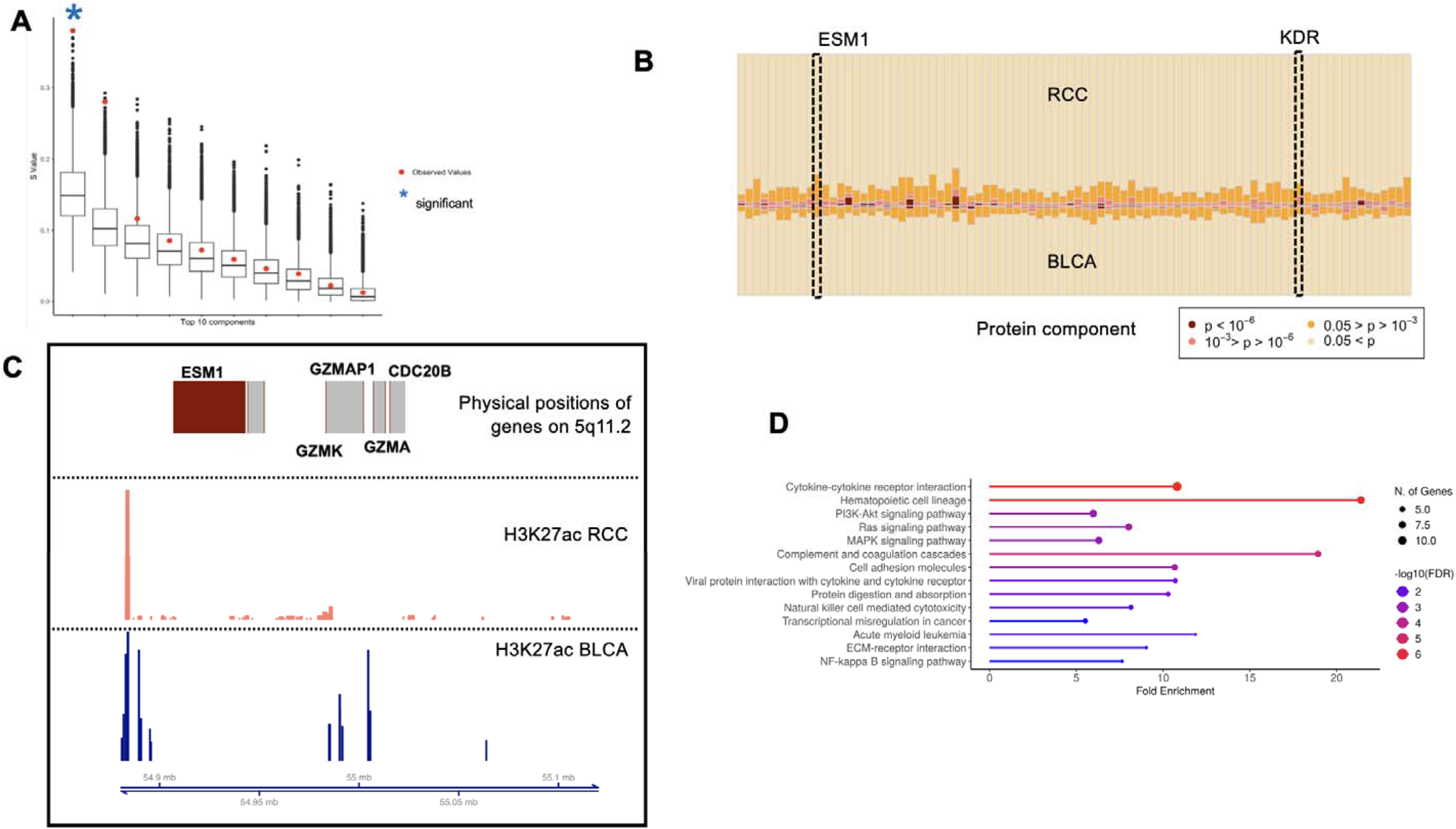
Results for genitourinary cancers. **(A)** Observed s-values (a measure of overall trans-associations captured by the protein components identified by TASTE; See **Methods**) in red dots against the expected distribution of s-values using a competitive hypothesis for the top 10 protein components. The significant associations are marked in blue asterisk. (**B**) Distribution of trans-pQTL p-values for each of the 88 proteins identified by TASTE in the protein component (horizontal axis), in terms of significance level for variants associated to RCC (upper part) and BLCA (lower part). KDR and ESM1 are highlighted having only nominally significant p-values. **(C)** Presence of H3K27ac marks in the cis-region of ESM1 in both RCC and BLCA tumors (See **Supplementary Methods**). (**D**) Pathway analysis of protein component identified by TASTE, showing the fold enrichment, number of overlapping proteins, and -log_10_(FDR) for significance.

In particular, 18 proteins had a strong trans-pQTL association (p-value < 1.1×10^-07^: Bonferroni threshold for all pairwise comparisons with 167 SNPs and 2,709 proteins) in both BLCA and RCC, of which 12 were identified by TASTE. Furthermore, among the 88 proteins identified, 38 proteins did not have a strong trans-pQTL association (p-value < 1.1×10^-07^: Bonferroni threshold for all pairwise comparisons with 167 SNPs and 2,709 proteins) with any one of the 167 SNPs indicating that these trans-associations were identified by aggregating multiple, possibly weaker associations, not identified by standard pairwise approach (**Figure 4B**). Notably, this included proteins like Vascular Endothelial Growth Factor Receptor 2 (KDR; minimum p-value = 1.4×10^-07^) which was only nominally associated (p-value < 0.05) to 10 RCC SNPs and 8 BLCA SNPs. KDR is primarily involved in angiogenesis, specifically through VHL–HIF–VEGF axis in RCC, and upregulation in BLCA. Querying OpenTargets platform^50^, we found that KDR has been the target in multiple ongoing or completed clinical trials including Pazopanib Hydrochloride in RCC and Sorafenib in BLCA, as vascular endothelial growth factor receptor inhibitor. Another such example is Endothelial Cell-Specific Molecule 1 (ESM1; p-value = 3.2×10^-07^), nominally associated to 14 RCC SNPs and 8 BLCA SNPs and can play an important role in tumor angiogenesis, inflammation, and aggressive tumor phenotypes in RCC and BLCA^51,52^. Using chromatin ChIP-seq data in RCC and BLCA tumors^53,54^ (See **Supplementary Methods**) we found that there are active open chromatin regions within 20Kb of the transcription start site of ESM1 (**Figure 4C**) indicating potential active regulation in these tumors.

Further in silico analysis of the 88 proteins identified by TASTE, revealed significant physical protein-protein interactions (p-value < 1×10^-16^) indicating that the proteins form important biological complexes potentially consequential to downstream cancer manifestation^55^. The selected proteins are also enriched in several known cancer processes like Ras signaling pathway (FDR = 7.2×10^-04^), MAPK signaling pathway (FDR =2.1×10^-03^) and PI3K/AKT pathway (FDR = 1.2×10^-03^) among others (**Figure 4D**; **Supplementary Table 4**), all of which have been implicated in both RCC and BLCA previously and in genitourinary cancers in general. In addition, the proteins are overrepresented in targets of transcription factors like TFAP2A (FDR= 1.8×10^-02^) and CTNNB1 (FDR = 3.5×10^-03^), which have been reported in context of several cancers (**Supplementary Table 4**).

We further explored publicly available cancer databases to assess the relevance of the TASTE-identified proteins for BLCA and RCC. Overall, among the 88 proteins identified, 4 proteins are reported to be tier-1 cancer drivers in COSMIC^46^ while 14 overall are candidate cancer drivers in NCG database^47^, including previously discussed proteins like KDR (**Supplementary Table 5**). Using gene expression data from matched tumors and normal in TCGA^56^ bladder urothelial carcinoma (BLCA) and clear cell renal carcinoma (KIRC), we found 65 (74%) were differentially expressed (FDR < 0.05) in at least one BLCA and KIRC (**Supplementary Table 6**), indicating potential relevance in respective tumors. Further, using immune infiltration inferred in TCGA^57^, we found that the presence of genetic variations in at least one of the 88 proteins was significantly correlated with lower CD4^+^ T cells in both BLCA and RCC tumors (**Supplementary Table 7**). Additionally, in publicly available CRISPR knock-out databases^58^, we identified 10 proteins were essential to cell proliferation for both and 26 proteins in at least one BLCA and RCC related cell line (**Supplementary Table 8**). Taken together, these highlight the potential importance of these proteins in context of both cancers.

## Discussions

While GWAS have successfully identified many genetic variants associated with different cancers, the comprehensive understanding of underlying biological mechanisms modulated by these associations remain elusive. Trans-genetic regulation by GWAS identified variants has been proposed as an effective avenue to identify potential target genes for different diseases including cancers. Coupled with the observation that related cancers can have overlapping molecular targets, this implies that joint analysis of trans-associations across multiple related cancers can highlight downstream targets that impact multiple cancers and can illuminate shared processes and pathways. Here, we have proposed a computationally efficient statistical method TASTE for this purpose, which aims to identify shared effects of omics, in particular proteomics, across groups of multiple related outcomes (cancers here). To our knowledge, there are no existing methods addresses this specific problem of identifying shared trans-associations across groups of similar cancers/outcomes.

Using *trans*-pQTL summary statistics (effect size, standard errors) of cancer related GWAS significant variants across the proteome, TASTE first estimates the joint or shared effects across multiple cancers in the group, through a matrix decomposition approach (TASTE-D). Subsequently, TASTE identifies the set of proteins (protein component) that contribute toward these shared effects (TASTE-E) and tests whether the identified protein-set represents a group-relevant trans-association pattern on a polygenic background of broader trans-associations expected in GWAS significant variants (TASTE-T). Simulations show that TASTE can reliably estimate the shared effects and identify the proteins contributing to the shared effects under several different scenarios. The accuracy of TASTE increases with increasing similarity between the constituents of the category as well as the number of GWAS significant variants within each group (cancer). Interestingly, we do not see a marked drop in performance even when the number of GWAS significant variants in each cancer differ greatly.

Application of TASTE to plasma proteomic data from UKB-PPP, on two groups of related cancers, identified several intriguing proteins that have shared effects on cancers in respective groups. For hematological cancers, we identified 161 such shared proteins overall, trans-associated to GWAS significant variants. Notably, we identified BCL2L1 protein, a member of the BCL2 family which has been reported to be associated with several hematological cancers^59,60^. This family of proteins is a fundamental regulator of the intrinsic apoptotic pathway which modulate cellular fate and their overexpression act as anti-apoptotic factors in many hematological malignancies. Additionally, the identified proteins were enriched among targets of MYC, an oncogene known to be particularly a driver in blood and immune related malignancies^31^. For genitourinary cancers, TASTE identified 8 proteins. This includes proteins like KDR and several other constituents of VEGF pathway which plays a crucial role in angiogenesis and immune response^29^, thereby influencing the development and progression of such cancers. We also find that the identified proteins shared across genitourinary cancers are overrepresented in PI3K-AKT, RAS and MAPK pathways all of which have central roles in formation, progression and drug resistance of these malignancies^61–63^. These analyses point out that the shared trans*-*association patterns detected by TASTE can highlight processes and mechanisms shared across similar groups of related cancers.

The protein component, which represents the set of selected trans-associated proteins shared across related cancers, is one of the key outputs of TASTE. In our analysis, for both the groups, we identify several proteins which do not have strong trans-associations with any cancer-related variants but are detected as a result of aggregation of several weaker associations across multiple distinct but related cancers. This highlights the statistical/informatic advantage that TASTE provides. Using a series of follow-up analyses for three different types of traits, we showed that the selected genes are often overrepresented in known relevant pathways, enriched in protein-protein interaction networks, contain targets for known transcription factors as well as manifest signatures in respective tumor tissues.

Although the estimation of shared effects matrix (TASTE-D) and protein components (TASTE-E) is computationally efficient, in the current implementation, the resampling-based competitive testing (TASTE-T) is computationally intensive. Further research is merited to develop analytical approximation techniques to reduce this computational burden. Since variable selection is a major goal in TASTE, we have performed sparse SVD via the L1 penalty due to its proven theoretical selection consistency^64^. However, in presence of correlation between protein levels, elastic net or fused lasso regularization approaches can also be effective alternatives^65^. In the future, the utility of such types of penalty functions in estimating the protein components merits further research.

Here we have analyzed plasma proteomic summary statistics reported by UKB-PPP primarily due to the substantial sample size yielding higher statistical power to identify trans-associations. While TASTE can be applied to QTL results from other tissues and cell lines, current sample sizes may be too limited to yield sufficient power. In general, it is expected that analysis in the cell type or tissue of origin for a given group of cancers would yield most relevant results. However, our analyses based on plasma did detect *trans*-association patterns that appear to have a broader biological basis in the context of the analyzed groups of cancers, from multiple independent lines of evidence. Nevertheless, it is likely that our analysis in plasma would capture immunoregulatory processes better and potentially miss *trans-*association patterns that are present only in specific tissues, cell types or/and dynamic stages. In the future, we will seek additional applications of TASTE in various types of newer omics databases to provide a more complete understanding of shared effects of molecular targets across an array of related diseases and outcomes.

There are several limitations of the proposed method and current analysis. First, in the current version TASTE, we begin with sets of (sentinel) SNPs associated with related cancers in a group, but we do not incorporate the underlying association effect sizes in the analysis. It is likely that incorporation of the GWAS effect sizes for the SNPs in TASTE could better identify and contextualize shared target proteins. Furthermore, we have only performed joint analysis of groups of cancers without explicitly quantifying their degree of overlap. Incorporating metrics for similarity of cancers, such as genetic correlation, can potentially be more informative for TASTE as well. Additionally, information on cis-genes and known functional/genomic annotations of genetic variants can further improve the power of the analysis.

In summary, we have developed a summary pQTL-based method, TASTE, to identify shared trans*-*associations patterns across multiple related diseases. TASTE is a powerful tool for identifying target protein networks that represent overlapping processes and mechanisms across similar cancers. Although, we have applied TASTE to large-scale publicly available proteomic data, extensions and adaptations of the method can easily be implemented for other relevant omics studies. In the future, applications of the methods to a variety of existing and new data on associations between genetic variants with high-throughput molecular traits can provide insights to biological mechanisms shared across related diseases.

## Data Availability

All data produced in the present work are contained in the manuscript and will be available in github.

## Acknowledgments

This work was supported by the Intramural Research Program of the National Cancer Institute, National Institutes of Health, US Department of Health and Human Services.

## Web Resources

GitHub repo for TASTE: https://github.com/diptavo/TASTE

UKB-PPP summary statistics: https://www.synapse.org/Synapse:syn51364943/wiki/622119

ShinyGO: https://bioinformatics.sdstate.edu/go/

GSCA: https://guolab.wchscu.Cn/GSCA/#/

gProfiler: https://biit.cs.ut.ee/gprofiler/gost

Crispr knock off data: https://depmap.org/portal/

PPI: https://string-db.org/

Cistrome DB: http://cistrome.org/db/

