## Supplementary Methods for "TASTE identifies shared proteomic effects on multiple related cancers"

**Supplementary Material**

TASTE-E: Algorithm to estimate protein components

**Input**: ${R, c}_{1}, c_{2}, \lambda_{v}, \gamma$ where $R$ is the matrix of estimated shared effects (See **Methods**), $\gamma$ is the step size and the rest are regularization parameters used in the algorithm.

Repeat the steps $r$ number of times.

1. Do until $c_{1}\leq\left\| \boldsymbol{v} \right\|_{0}\leq c_{2}; \left\| \boldsymbol{v} \right\|_{1}\leq\lambda_{v}$

- $\left( \boldsymbol{v} \right)\boldsymbol{=}\boldsymbol{argmax}_{\left\| \boldsymbol{v} \right\|_{2}\boldsymbol{=1}}\boldsymbol{v}^{\boldsymbol{T}}\boldsymbol{R}^{\boldsymbol{T}}\boldsymbol{Rv}$
- $s={{(1}^{T}Rv)}^{2}$
- Calculate $\left\| \boldsymbol{v} \right\|_{0}$
- $\lambda_{v}=\lambda_{v}+\gamma$

2. From $\boldsymbol{v}$ obtained above, store the indices of components which have non-zero entries (corresponding proteins will be selected), and the corresponding s-value.
3. $R^{*}=R-Rvv^{T}$
4. $R=R_{sub}$ where $R_{sub}$ is obtained by removing the columns that have non-zero loadings in sparse vector $\boldsymbol{v}$ **.**

**Output**: an estimated sparse vector $\boldsymbol{v}$, denoting the protein component (See **Methods**). The non-zero elements of the protein component $\boldsymbol{v}$ correspond to the proteins selected.

We estimate as many protein-components possible by iterating until the columns (proteins) of $\boldsymbol{R}$ are exhausted, ensuring no overlap between proteins selected in each component. In our implementation we set, $c_{1}=30, c_{2}=100$ and $\gamma$ = 0.1.

Simulations: Algorithm to generate data for evaluating the performance of TASTE

**Input**: ${p, r, g}_{1}, g_{2}, r_{1},r_{2}$ are parameters denoting the total number of proteins, rank of shared effect matrix, number of genetic variants associated to each disease, ranks of disease specific effect matrix (See **Simulations** for details).

Repeat the steps $r$ number of times.

1. Generate $U_{i},L_{i},Q_{i}, \varepsilon_{i}$ and $V , i=1,2$such that:

- The rows of $U_{i}$ is $g_{i}$ independent observations from $N(0,I_{r})$
- $V^{T}=[\beta I_{r}, 0_{r\times(p-r)}]$
- $Q_{1}^{r_{1}\times p}=[0_{r_{1}\times r}, \frac{\beta}{2}I_{r_{1}}, 0_{r_{1}\times(p-r-r_{1})}]$
- $Q_{2}^{r_{2}\times p}=[0_{{r_{2}\times(r+r}_{1})}, \frac{\beta}{2}I_{r_{2}}, 0_{r_{2}\times(p-r-r_{1}-r_{2})}]$
- $\varepsilon_{i}$ is $g_{i}$ independent observations from $N(0,I_{p})$

2. $Z_{i}=R_{i}+S_{i}+\varepsilon_{i}$ where $R_{i}= U_{i}V$, $S_{i}= L_{i}Q_{i}$
3. Apply TASTE-D on $Z$ to extract joint rank and joint effect matrices $R.$
4. Apply TASTE-E $R$ to estimate the protein components (select columns contributing to the shared effect matrix)
5. The estimated joint rank is noted, and sensitivity and specificity are calculated.

**Output**: Matrices of z-values $Z_{i}$ with $g_{i}$ rows (corresponding to associated genetic variants) and $p$ columns (corresponding to proteins) for $i=1,2$. Each $Z_{i}$ represents to the z-value matrix for disease/cancer $i$ among the group of two related diseases/cancers.

**In silico analysis.**

A. *Protein-protein interactions*: To evaluate whether the proteins selected in the protein components manifest more frequent physical interaction than expected by chance, we calculated the protein-protein interaction enrichment statistics using the STRING Db software (See **Web Resources**).

B. *Pathway enrichment*: To evaluate the overrepresentation of the proteins selected in the protein components in specific pathways or biological processes than expected by chance alone, we performed pathway enrichment. For this we used ShinyGO (See **Web Resources**) which assembled pathways, gene-sets and biological processes across several public databases spanning KEGG, GO, REACTOME and others. The corresponding web tool performs a hypergeometric test of enrichment.

C. *Cancer genes*: For proteins selected in the protein components, we checked whether these were included in COSMIC database as known cancer driver genes or in NCG database as candidate cancer driver genes (See **Web Resources**).

D. *TCGA analysis*: Multiple analyses were conducted using the molecular resources provided by the cancer genome atlas (TCGA) project for the cancers analyzed (See **Results**). These included differential gene expression analysis and analysis of immune infiltration. The analyses were conducted using the GSCA platform (See **Web Resources**) which provides several inferred molecular resources using TCGA data including immune infiltration estimated by ImmuneCellAI.

E. *chromatin ChIP-seq*: We curated H3K27ac ChIP-seq data from in BLCA cell lines from Cistrome DB and from the data reported by Nassar et al for RCC tumors.
